# FFT Power Relationships Applied to EEG Signal Analysis: A Meeting between Visual Analysis of EEG and Its Quantification

**DOI:** 10.1101/2025.03.14.25323563

**Authors:** Juan M. Díaz López, Jose Curetti, Vanesa B. Meinardi, Hugo Díaz Farjreldines, Carina Boyallian

## Abstract

**Objective:** This study presents a novel computational approach for analyzing electroencephalogram (EEG) signals, focusing on the distribution and variability of energy in different frequency bands. The proposed method, FFT Weed Plot, systematically encodes EEG spectral information into structured metrics that facilitate quantitative analysis.

**Methods:** The methodology employs Fast Fourier Transform (FFT) to compute the Power Spectral Density (PSD) of EEG signals. A novel encoding technique transforms frequency band distributions into six-entry vectors, referred to as “words,” which serve as the basis for three key metrics: a scalar value 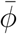 a vector 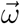, and a matrix *H*. These metrics are evaluated using a dataset comprising EEG recordings from 30 healthy individuals and 15 patients with epilepsy. Machine learning classifiers are then applied to assess the discriminatory power of the proposed features.

**Results:** The classification models achieved a 95.55% accuracy, 93.33% sensitivity, and 96.67% specificity, demonstrating the robustness of the proposed metrics in distinguishing between control and epileptic EEGs.

**Conclusions:** The FFT Weed Plot method provides a novel approach for EEG signal quantification, improving the systematization of spectral analysis in neurophysiological studies. The metrics developed could serve as quantitative descriptors for automated EEG interpretation, offering potential applications in clinical and research settings.

**Highlights:**

- From frequency domain analysis to information and probability theory, new ways of encoding information.
- A step towards the systematization and automation of medical EEG reading.
- New global metrics for the description of the energy of an EEG recording and their applications in machine learning.
- The FFT Weed Plot method, We present a new, reproducible, robust and clinically designed method to improve the objectivity of medical practice and research in neurophysiology.

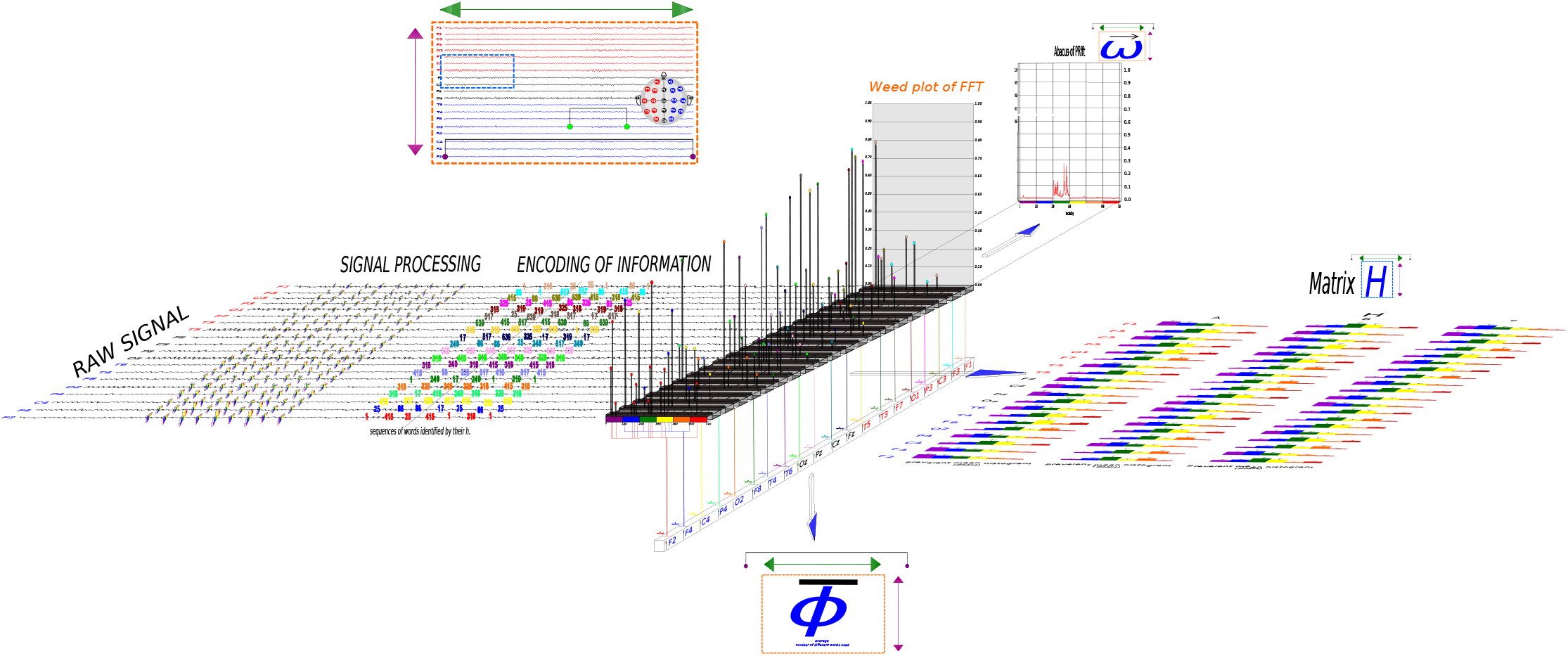

## 1. Introduction

An electroencephalogram (EEG) is a graphical representation of neural activity that is registered using electrodes placed on different parts of the scalp. Each electrode mainly samples the synaptic activity that occurs in the superficial layers of the brain cortex. In clinical neurophysiology, the EEG is an instrument for observing and monitoring the electrophysiological activity of the brain for evolutionary and prognostic diagnosis.

Usually, clinical interpretations of an EEG are achieved by associating pathological features with certain pattern recognition during visual inspection Ebersole and Pedley [15],Tong and Thankor [52]. Although this traditional analysis is quite useful, it is subjective and does not easily allow for any systematization Ré et al. [39], Nascimento et al. [32].

The interpretation of an EEG has two stages: descriptive and inferential. The first stage includes all the descriptive aspects of the signals, namely analyzing the representative graphoelements, the inter-hemispheric symmetry, the synchronicity between signals, and the distribution and variability of the energy of the signal Tsuchida et al. [53], Hirsch et al. [20]. On the other hand, in the second stage, contextual information, such as age, consciousness state, physiological condition, duration, and psychopharmacological prescription before EEG recording, is considered Niedermeyer et al. [33]. At this point, inference occurs and physicians can provide a report on neurophysiology diagnosis. Sometimes this report contains both interpretations, but a clear inferential interpretation is not always possible. This is because the signs present in the EEG are non-specific to a unique pathology and also these signs can be considered as normal or abnormal waves depending on the stages of neurodevelopment or consciousness state. This complexity underlies EEG interpretation [15, 58]. For all of the above reasons, EEG analysis has traditionally relied on visual inspection by trained neurophysiologists, a process which, although effective, requires many years of medical training, lacks standardization and is inherently subjective.

In recent years, quantitative EEG (qEEG) analysis has gained attention, the quantitative analysis of an EEG recording was mainly based on the use of classical techniques of signal processing Harmony [19], Tong and Thankor [52], Oppenheim et al. [35], Blanco et al. [7], Sanei and Chambers [46]. Methods related to wavelets Rosso et al. [44], Bethany Gosala [6], Yang et al. [60], [2, 43], which can be thought of as a generalization of Fourier Transform Anuragi et al. [4], Mehla et al. [31], Al-Salman et al. [3], and others related to information theory, using Lempel-Ziv complexity [55, 13, 23, 12, 30], entropy Cataldo et al. [11], Guisande et al. [18], Gancio et al. [16], and Jensen-Shanon divergence Pereyra et al. [36], Lamberti and Majtey [24], Ré et al. [40], Mateos et al. [29], are proposed to find information in EEG signals Vicente et al. [54], Bossomaier et al. [10], Restrepo et al. [41].

In addition, the rise of artificial intelligence has led to the development of new methods for classifying EEG recordings Tjepkema-Cloostermans et al. [51], White et al. [56]. These methods extract information from the signals by dividing them into EEG bands Huang and Ling [21] and applying neural networks to classify the extracted characteristics [49, 26, 59].

However, applying these methods to daily physicians’ practice is not always easy. In some cases, physicians may find it challenging to use these methods due to their focus on mathematical and physical tools that are difficult to introduce. In other cases, the methods may not capture the unique features and singularities of physicians’ EEG interpretation. These abilities are often hidden in the expertise and experience of clinical physicians.

In this work, we have developed and applied a new method for quantifying, analyzing, and comparing EEG recordings. Our approach approximates the quantitative analysis (qEEG) to the visual analysis (vEEG) of EEG by formalizing the way physicians visually inspect power proportions across bands. Our goal is to understand the dynamics of energy variability in EEG recordings over time and among all electroencephalographic bands, not just the band with the highest power. The method developed in this article focuses on the process of medical EEG reading and its descriptive interpretation for diagnostic purposes. Here we introduce a novel computational method, the FFT Weed Plot, which encodes EEG spectral information of Power Spectral Density (PSD) analysis into structured representations that enhance the interpretation of EEG power distributions. The method systematically quantifies the relative power proportions across EEG bands using lexicographic ordering, yielding three primary metrics: 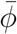 (scalar): Average vocabulary complexity of EEG spectral patterns, a vector 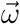 (vector): Normalized prevalence of spectral words across EEG channels., and *H* (matrix): A representation of energy distribution dynamics across EEG bands.

To evaluate the effectiveness of these metrics, we applied the method to a dataset consisting of 30 control subjects and 15 patients with epilepsy. Machine learning classifiers were employed to assess their discriminative power, achieving an accuracy of 95.55% in distinguishing between normal and pathological EEGs.

In addition, this method could provide a population-based frame of reference to contrast the dynamics of a single EEG recording in a single case study. This is part of our ongoing work The structure of the article is the following: Section 2: describes the proposed methodology, including the FFT-based feature extraction and coding process and the design of the experimental setup. Section 3 analyses the results of the study, based on the different machine learning methods applied and the results of the individual and joint behavior of the three metrics proposed to discriminate between the two groups. In Section 4, our results are compared with those obtained by similar studies, followed by a discussion of clinical and research implications. Section 5 summarizes the study’s key findings, as well as suggests further research using this methodology.

Summarizing, this study proposes a novel EEG analysis method that formalizes the visual inspection process by encoding power spectral distributions into structured quantifiers. Our hypothesis is that these FFT-based power relations can effectively differentiate normal EEG recordings from those with pathological features, bridging the gap between subjective clinical interpretation and quantitative signal analysis. This approach aims to enhance standardization in neurophysiological diagnostics while maintaining interpretability for clinicians.

## 2. Materials and Methods

### 2.1. Preliminaries

The electrical brain activity produces a continuous time analog signal *X*_*a*_ (t). Nevertheless, studies of brain activity using digital electroencephalography transform the analog signal to a digital one with discrete values for discrete time. This sampling produces a related signal which is a set of discrete points. The interval of time between samples has to be regular. We name it *T*_*s*_. Thus we have

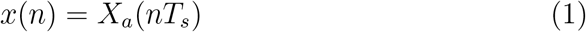

with n ∈ ℤ_≥0_ and where x(n) is obtained by taking a sample value of the analog signal *X*_*a*_ every *T*_*s*_ seconds. The time interval *T*_*s*_ between successive samples is called *the sampling period* or *the sampling interval*. The *sampling rate or sampling frecuency* is *f*_*s*_ = 1/*T*_*s*_ samples per second or Hz.

The sequence of N numbers x(0), …, x(*N*− 1) obtained sampling our analog signal is transformed to a sequence of complex numbers X(0), …, X(*N* − 1) through the Discrete Fourier Transform (DFT), Bloomfield [8] as follows:

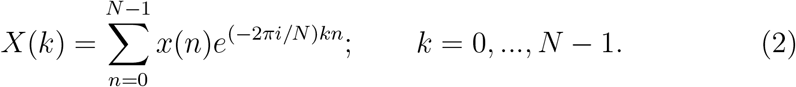

The Discrete Fourier Transform changes from the time domain to the frequency domain. The Parseval theorem states

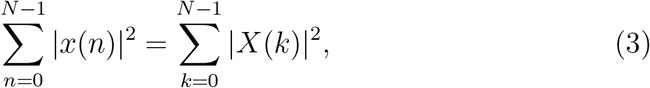

so, the energy of the signal is the same as the energy of its Discrete Fourier Transform Wiener [57]. We denote this energy by *E*.

For each frequency k, we call *Power Spectral Density* (PSD) to:

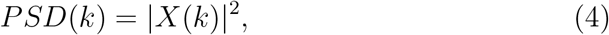

and so the

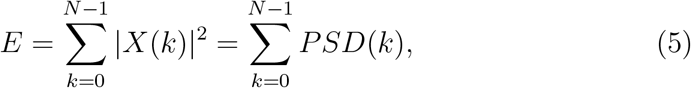

that is, the Energy is the sum of all Power Spectral Densities.

In Clinical Neurophysiology, electrical waves are studied in frequency bands, each with its own range of frequencies. However, only frequencies that comply with the Nyquist criterion can be studied. To sample an analog signal with a frequency *f*_*s*_ and avoid aliasing [35], the highest frequency that can be considered should be equal to or less than *f*_*s*_/2. This condition is known as the *Nyquist criterion*, which states that the sample frequency *f*_*s*_ must be at least twice the bandwidth of the signal [34, 48, 25].

Each hardware has its own sampling rate to transform analog signals into digital ones. In other words, not all hardware samplings allow for the study of all EEG bands. Therefore, it is necessary to take the Nyquist criterion into account when choosing the bands to be studied. Since we used equipment with a sampling rate of 65 Hz in this work, the electroencephalographic bands considered are:

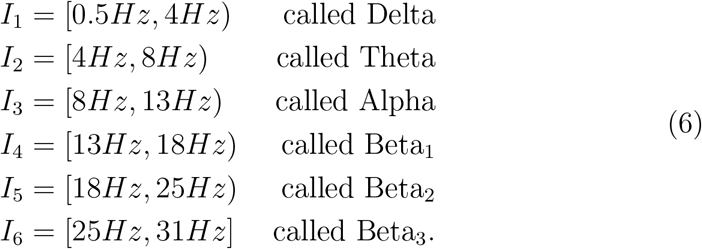

For each value of b ranging from 1 to 6, we define the “Accumulated Power per Band” as follows:

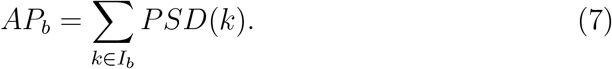

This definition produces a six-entry vector called the “Accumulated Power per Band Vector,” denoted by 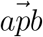. It stores in each entry b the percentage of the total energy contained in the corresponding I_b_ band. A practical way to present clinically relevant information about the accumulated power per band is to use the “Relative Power Histogram” (RPH), this is a step function that assigns to each interval I_b_ the percentage of the total energy contained in its band. Figure 1 shows an example of an RPH.

**Figure 1:**
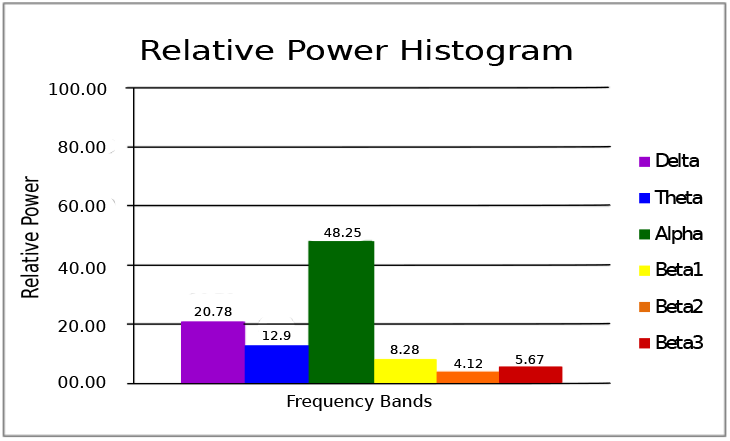
Each electroencephalographic band corresponds to a percentage value that represents its relative power.

In neurophysiological clinical practice, physicians refer to ‘the dominant band’ as the electroencephalographic band with the highest percentage in the RPH. Namely, it is the one with the highest energy and the most accumulated power.

Our method considers the physical meaning of the RPH and how physicians use this information in their clinical practice.

### 2.2. Method: The FFT Weed Plot

#### 2.2.1. The Words or Power Relations

In this part, we will introduce a novel quantification method for EEG that systematizes the way physicians read them.

As a key element of our method, we define a word 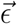 to be a vector with six inputs, each of which corresponds to a different frequency band in the EEG signal. Specifically, ϵ_1_ corresponds to the Delta band, ϵ_2_ corresponds to the Theta band, and so on, up to ϵ_6_, which corresponds to the Beta 3 band.

Given an 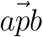 or its corresponding RPH, we define the *Power Relation* as the word 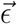, where each entry encodes the relative decreasing order of the attached band according to its energy in the RPH. To illustrate how to calculate the Power Relation, consider Figure 1: the decreasing order of the bands according to their energy is Alpha, Delta, T heta, Beta_1_, Beta_3_, and Beta_2_. Thus, 1 corresponds to Alpha or ϵ_3_, 2 to Delta or ϵ_1_, 3 to T heta or ϵ_2_, 4 to Beta_1_ or ϵ_4_, 5 to Beta_3_ or ϵ_6_, and 6 to Beta_2_ or ϵ_5_, yielding 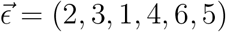.

An important point to note is that many different 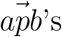 may have the same power relation s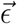. This means that our method can capture similarities between EEG signals that may not be immediately apparent from their raw data, since different signals can have the same underlying power relation.

We define the *vocabulary* as the set of all 6! = 720 possible power relations. In this vocabulary, we define the following lexicographic order: Given two words 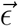 and 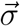, we say that 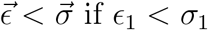 if ϵ_1_ < σ_1_ or if ϵ_i_ = σ_i_ for i = 1, …, k with k < 6 and ϵ_k+1_ < σ_k+1_.

Let h be the ordinal of a given word in the vocabulary. Since we have a complete order, we name the word by its h-word, h-power relation, or simply write 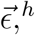.

The dominant band is the band attached to the entry where 1 is placed in 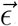. In our example, 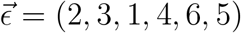, and 1 is placed in the entry ϵ_3_ attached to the Alpha-band. Thus, we say that the dominant band is Alpha. Let’s check another example. Take 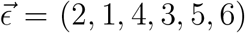. Here, the dominant band is the T heta-band since the value 1 is located in the second input of ϵ.

This way of encoding the information obtained from the RPH differs from physicians’ usual readings. They consider only the two bands with more energy, but ours is a new perspective, where the energy of all electrophysiological bands are simultaneously taken into account.

#### 2.2.2. Processing the raw signal

At this point, it is necessary to know the data structure, the number of EEG recording channels, the sample rate, and the duration of the recording, among other things.

Similar to Permutation Entropy from Information Theory or Spectogram Multitapper, our method uses a sliding rectangular window through the signal [28, 37]. In our case, we tested the window length (Δ) and step size (τ) for Δ = 4 and Δ = 2 seconds with τ = 0.25 and τ = 0.50 seconds. However, we ultimately chose a window length of Δ = 2 seconds and a step size of τ = 0.25 seconds to capture brief events in the EEG activity.

For a given EEG signal and each channel, we proceed as follows:

1. Place the rectangular sliding window of length Δ at the beginning of our raw signal. In our case, Δ = 2 seconds, so we have the first 130 data points of our signal. Generally, we set the position of the window as the place of its last data point. For this first window, it is at 130. Then,
  - Apply the FFT to this window and compute the Accumulated Power AP_b_ for each band I_b_ with b = 1, *· · ·*, 6.
  - Finally, encode that information in a word named 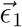, as described at the beginning of the section, and let h_1_ be its corresponding order in the vocabulary. We also set a counter f_1_ = 1, which will count new words as they appear while we slide the window through the signal.
2. Next, we slide the window forward, in our case by τ = 0.25 seconds, which means we move forward 17 data points. So, the position of our window is now at 146. Note that the new window includes data points 16 to 146. As in our first window, we apply FFT, compute the PSD for each band, 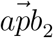, RPH_2_, and finally, we have the corresponding word 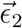 and its order h_2_ for this window. At this point, we set *f*_2_ = 1 if *h*_1_ = *h*_2_ or *f*_2_ = 2 otherwise (see Figure 3).
3. Then, we keep moving our sliding window forward by τ = 0.25 seconds, attaching a 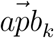, a word 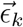, and its order *h*_k_ to each k-window until the sliding window position reaches the last data point of our signal. Note that the subscript k refers to the corresponding step in the procedure, and *f*_k_ gives the number of different new words that appeared until the *k*-step.

In the end, for each EEG channel, we have three sets: the set of all 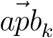 or their graphical representations, the RPH_k_, the set of all words 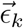, and their corresponding orders *h*_*k*_ in the vocabulary, which encode our original signal, and the counter f, which gives us the number of different words that appeared (see Figure 2).

**Figure 2:**
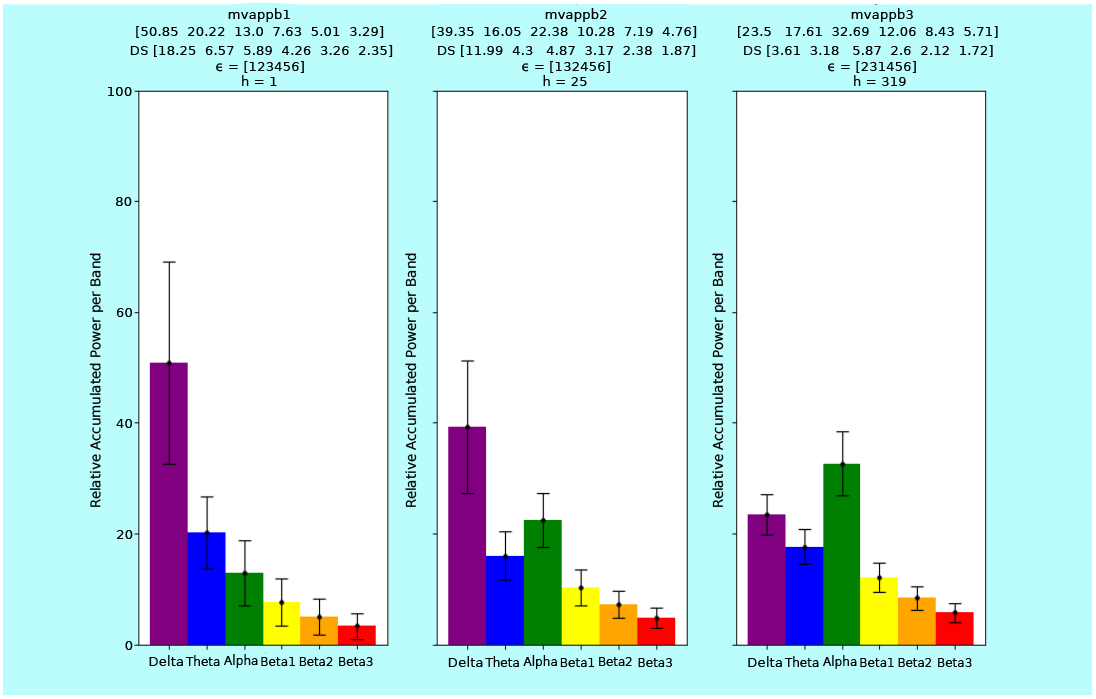
Each histogram shows its ϵ and the corresponding h.

**Figure 3:**
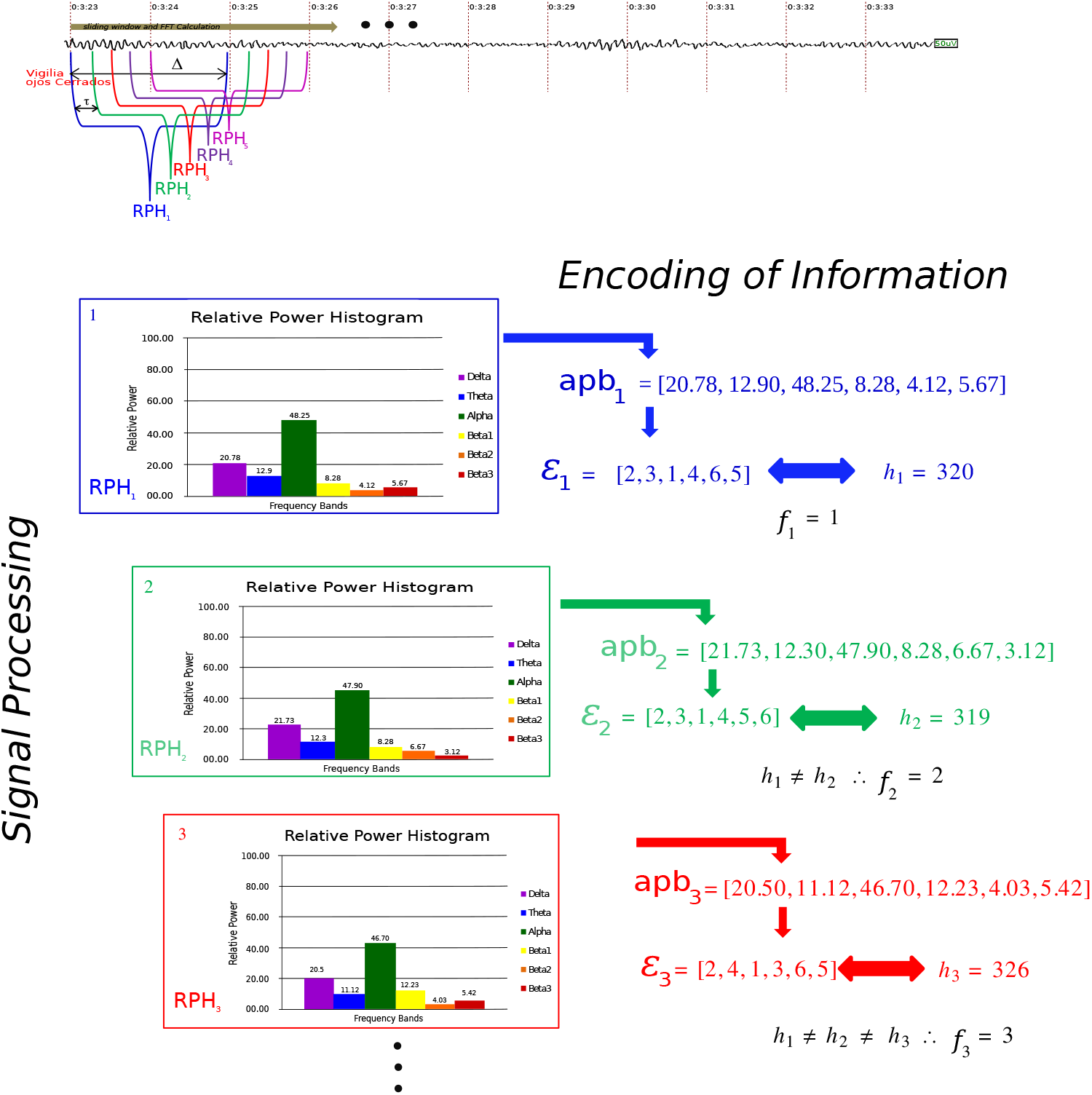
For each window of width Δ, the data is transformed from the time domain to the frequency domain using the FFT. The accumulated power per band is then calculated. This information is visualized in its Relative Power Histogram (RPH). The data is encoded in a vector 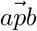, and the corresponding Power Relation and lexicographic order h are determined. The process is repeated by advancing one step τ, iterating until the end of the time series.

We call the *word pool of the channel* c, denoted by W_c_, the set of words obtained for this given EEG channel, and *Z*_c_ the set of all 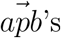. We suggest that the number of words in *W*_c_ or vectors in *Z*_c_ which corresponds to the total number of windows, related to the length of the signal, should be close to an order of magnitude greater than the size of the vocabulary.

#### 2.2.3. The output: Construction of the metrics

To construct the metrics we will need to give an order of the EEG channels. In our case we are dealing with 20 channels ordered from 1 to 20 in the following way, (see Figure 4):

**Figure 4:**
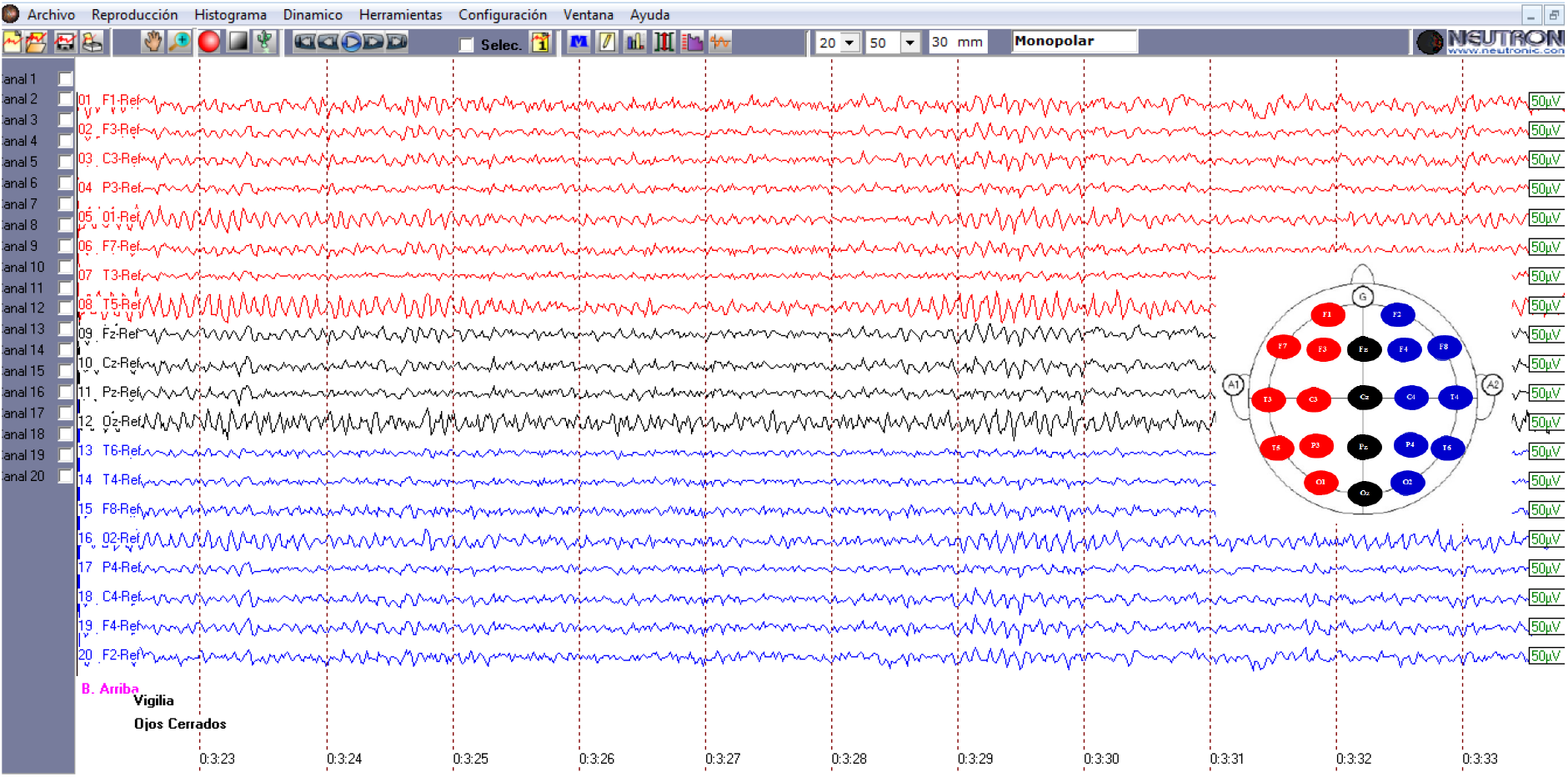
In our case, we are working with 20 channels, ordered from 1 to 20 based on how the monopolar channels were arranged in the hardware used. The equipment used was a Neutronic 24-channel polysomnograph, model ME-2400, with bimastoid references [47].

**Figure 5:**
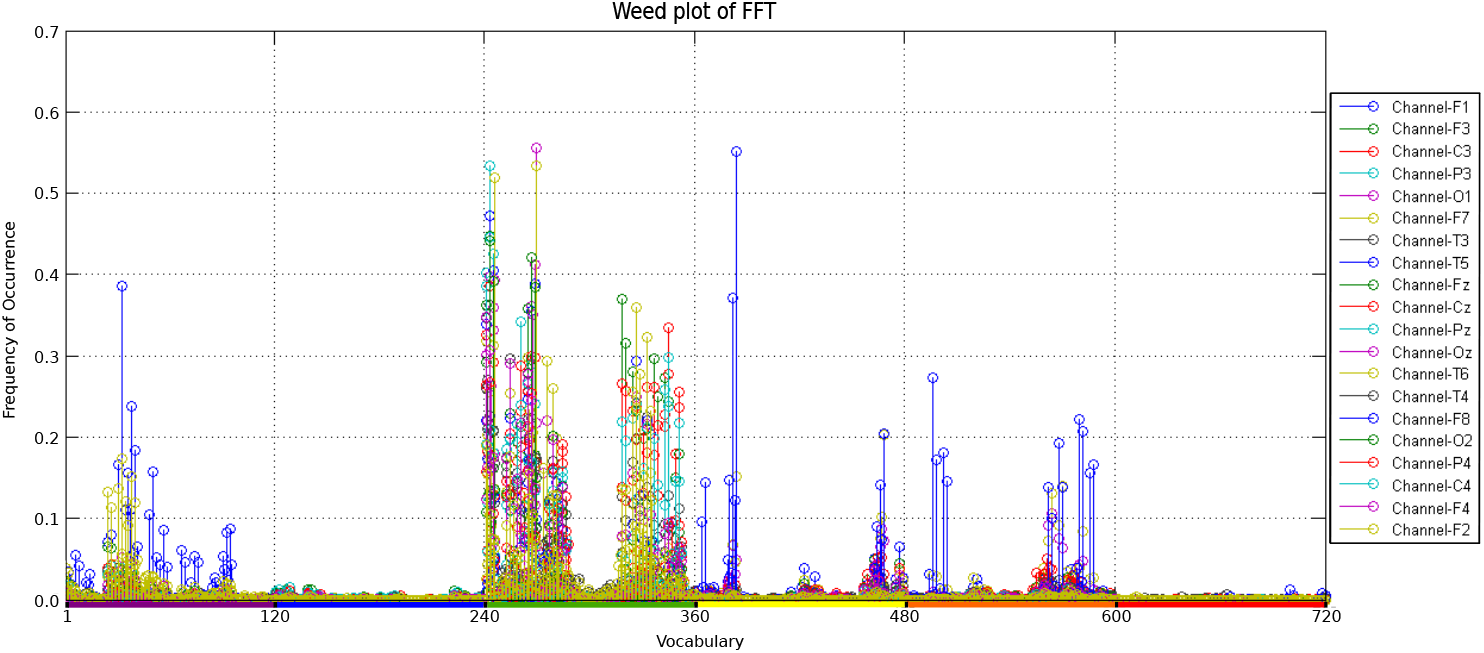
The weed plot of the FFT is a graphical representation of the matrix M. The colored segments below the X-axis indicate changes in the highest energy or dominant power among the ordinal ranks of the words in our vocabulary.

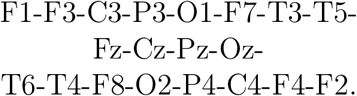

Our recordings for the control group were made with a Neutronic brand, 24channel polysomnograph, model ME-2400 with bimastoid references. This hardware ordered the channels in this way. Let’s introduce the following metrics.

1. 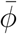 is the average number of different words used, and it can be calculated by counting the number of different words that appeared in each vocabulary W_c_, namely the value of the counter *f*^*c*^, which corresponds to the value *f*_*k*_ of the last window slid through the signal. The average of these values over all channels c gives us the average number of different words used during the whole recording.
2. The Frequency of Occurrence Matrix M and the abacus of PRfft, 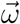.

We are also interested in the number of times each word appeared in W_c_. To reorganize the encoding data, a vector V_c_ will be created for each channel with 720 entries, one for each word in our vocabulary ordered according to the lexicographic order given above. The first entry corresponds to the word 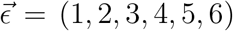, the second to 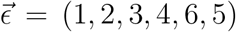, the third to 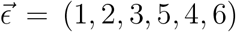, and so on. The frequency of occurrence of the corresponding word in W_c_ is assigned to each entry, and the vector is then normalized by its Euclidean norm. We call this vector the (normalized) *prevalence vector for channel* c.

To manage all EEG channels, we rearrange the prevalence vectors into the “frequency of occurrence matrix” *M*, which is a 720 × 20 matrix with columns that are vectors of normalized prevalence *V*_*c*_. Note that in this way, we can check the normalized frequency of occurrence of a given word in each channel by reading each row.

To plot each *V*_*c*_, we select a different color for each channel and set the numbers 1 to 720 on the x-axis to represent the ordinals of the words in our vocabulary. We plot the normalized frequency of occurrence of the corresponding word in this channel on the y-axis (see Figure 1). We call this the “Weed plot of FFT”.

To obtain global information for each EEG recording, we compute the mean occurrence of each word in the whole EEG by taking the mean of each row of the frequency matrix M. This yields a new 720-entry vector that we call 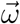. As with each *V*_*c*_, we plot 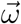 and call it the “Abacus of PRfft” (see Figure 6).

**Figure 6:**
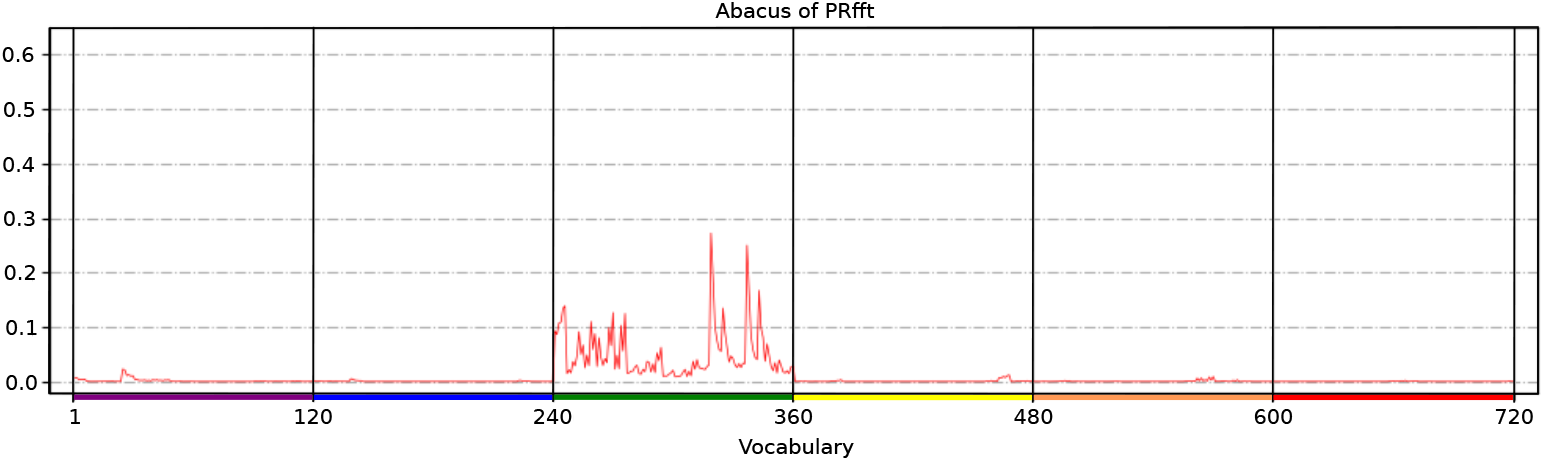
The abacus of PRfft is the graphic representation of 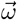

- The average cumulative percentage power per band matrix *H*

We go back to our matrix M, and we choose the three highest values of each column. Based on their position in the column, we can recover the three most prevalent words in the corresponding channel. We denote such words by A_c_, B_c_, and C_c_, where A stands for the word with the highest frequency of occurrence, B for the following one, and C for the word with the lowest among the three most prevalent ones in a given channel c.

Recall that *Z*_*c*_ is the set of all 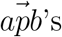 of channel *c*, and for a given word, you may have many 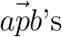 that correspond to it.

We recover from 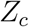 the sequence 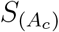 of all 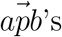 that share the same power relation with the word A_c_. We do the same for B_c_ and C_c_, recovering the corresponding sequences 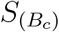 and 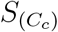.

Let *X* denote *A, B*, or *C*. Given a sequence 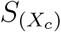, we arrange its elements in a six-column array 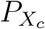 so that its columns correspond to the values of cumulative percentage power of the corresponding band.

Then, we compute the mean of each column of 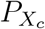, and we rearrange this information in a new six-entry vector called the mean percentage vector of cumulative power per band, denoted by 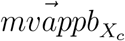 corresponding to *X*_*c*_ (see Figure 2). Thus, we have three such vectors, one for *A*_*c*_, *B*_*c*_, and *C*_*c*_. Since we are dealing with 20 channels c, at the end, we will have 60 such vectors that we rearrange in a new 60 *×* 6 matrix *H* as follows:

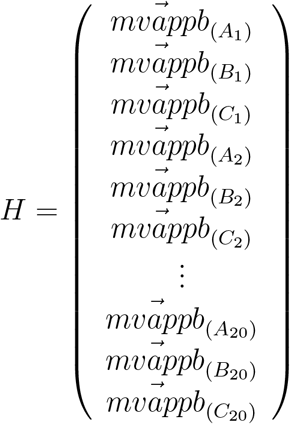

We represent each 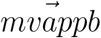, i.e., each row of H, as a step function that associates each band interval I_b_, see equation 7, in the frequency domain with the value of its corresponding entry. This function is called the prevalent mean histogram (PMH) (see Figure 2). Therefore, we have 60 PMHs, three for each EEG channel.

Note that each 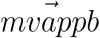 and its graphic representation PMH recover the distribution of the size of the percentage energy per band of the corresponding three most prevalent words of the corresponding EEG channel. The H matrix is the third relevant metric of the method.(see Figure 7)

**Figure 7:**
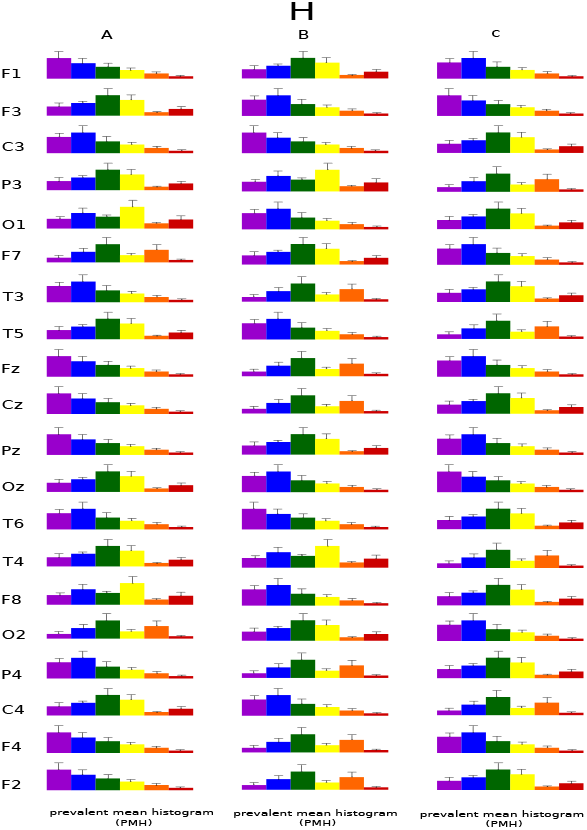
The graphic representation of matrix H

##### Summarizing, for each EEG, we have the following metrics

- The average vocabulary used by the EEG, 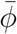.
- 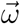 whose values are the average frequencies of occurrences of each word in all channels, and its plot, the Abacus, which allows us to see in a single image the behavior of the total energy of the EEG recording.
- The matrix *H*, which recovers the distribution of the sizes of the percentage energy per band of the corresponding three most prevalent words of the corresponding EEG channel.

##### Synopsis of the Weed Plot method. See Figure 8

These are the metrics used in this work. However, more metrics can be defined with this method. For instance, the dynamic abacus, which is the 20-column matrix whose columns are given by the vectors of the ordinals of the words corresponding to the channel c, or the growth curve, where we arrange the vectors F_c_ with entries f_k_ for each channel c as columns of our matrix. We are not dealing with them in this article, and they are part of the work in progress.

**Figure 8:**
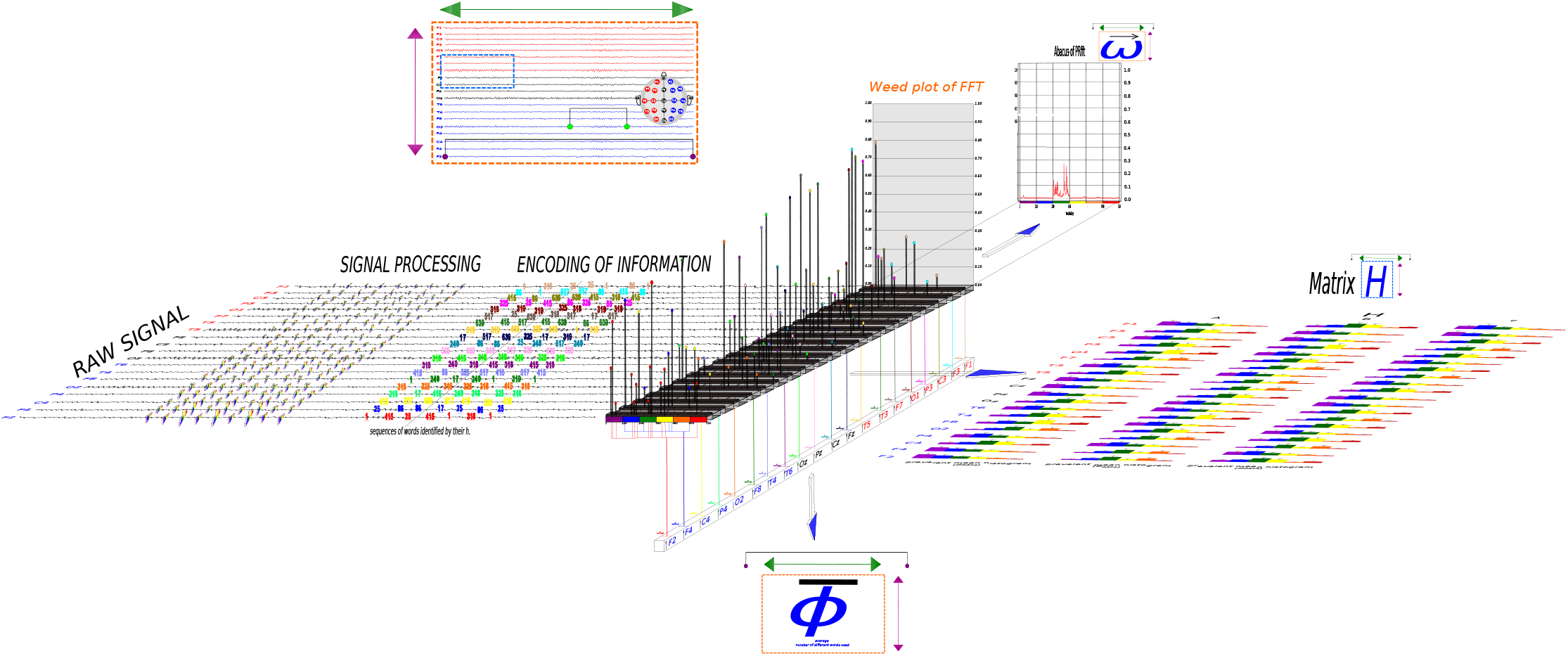
Synopsis: The medical interpretation of an EEG involves at least three key metacognitive processes: a) Direction of reading: Vertical and horizontal scanning, represented in the recording image by purple and green arrows. b) Visuoperceptive field: Represented by purple arcs for the macrotemporal scale (minutes or hours) and a green arc for the microtemporal scale (seconds). c) Focus of observation: Depicted by dashedline squares. A light blue rectangle indicates focal observation, while an orange rectangle represents global observation. In this synopsis of the Weed Plot method, we demonstrate how the three quantifiers are constructed to incorporate these metacognitive processes in both their creation and interpretation. The graph illustrates the process, beginning with the transformation of the raw EEG signal in the time domain using a sliding window and applying the FFT. This produces Relative Power Histograms (RPHs) for each EEG channel. The RPH data is encoded into sequences of words for each channel, which are subsequently represented by their lexicographic order, h. The complete EEG recording is then reorganized into the Weed Plot, from which the three quantifiers are derived. The symbols associated with the quantifiers—ϕ (number),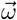 (vector), and H (matrix)—relate to the metacognitive aspects of EEG interpretation outlined above. This framework integrates the direction of reading, the visuoperceptive field, and the focus of observation, offering a comprehensive approach to EEG analysis.

### 2.3. Application of the Weed Plot method. A small experimental design

To create a proof of concept for the method, we designed a small experiment based on the contrast between two groups of EEG recordings.

The control group, referred to as group A, consisted of 30 individuals with a normal EEG report confirmed by a neurophysiologist. The inclusion criteria were: a) women and men aged between 20-50 years old; b) individuals with at least 7 years of formal education; c) individuals without any neurological or psychiatric medical history and normal values on the Beck Depression Inventory [9, 17] and State-Trait Anxiety Inventory, forms X and Y [50]; d) individuals with a normal EEG report based on St. Louis EK and Frey LC’s criteria of normality [27]; and, e) individuals who had accepted and signed the informed consent.

The exclusion criteria included: a) adults over 50 or under 20 years of age; b) individuals who have not been literate; c) individuals with a clinical history record indicating any issue; d) individuals who consume psychotropic drugs or drugs with effects on the Central Nervous System; e) individuals who have an abusive consumption of psychoactive substances such as cocaine, marijuana, alcohol, or others; f) individuals who consumed moderate or minimal alcohol in the 48 hours before the complete research evaluation; g) individuals who scored above 9 points on the administered BDI scale; h) individuals who obtained values above the 70th percentile for anxiety symptoms on the STAI X scale for anxiety; i) individuals whose electroencephalographic studies showed paroxysmal activity or any other detected functional abnormality; and, j) individuals who did not agree to sign the Informed Consent.

The research protocol was approved by the ethics committee of Hospital Córdoba, Córdoba, Argentina and registered in the Ministry of Health of the Córdoba Province on 20-03-2017 under number 3170.

Each of these EEGs corresponds to the basal electrophysiological condition, i.e., awake with closed eyes. The recordings were made using a Neutronic brand 24-channel polysomnography, model ME-2400 with bimastoid references. The average length of each recording is 30 minutes, and they were sampled at 65Hz.

The second group, referred to as our “single case study” group (SCS), comprises n=15 recordings of patients with epilepsy of different causes, who presented generalized tonic-clonic seizures. This data was obtained from “The TUH EEG Epilepsy Corpus (TUEP),” which is a subset of TUEG containing 100 subjects with epilepsy and 100 subjects without epilepsy, as determined by a certified neurologist. The data was developed in collaboration with several partners, including NIH.

All the pathological recordings we considered for this work correspond to intercritical EEGs in epileptic patients [5], and they were down sampled to 62Hz after decimating the raw signal, which had an original sampling rate of 250Hz. They have an average duration of 30 minutes each.

All EEG’s were processed identically applying our method so we have a set of

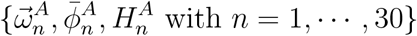

for control group *A*, and

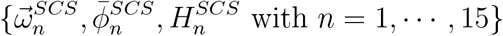

for the single case study group.

We arrange the 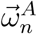 as columns in a 30 × 720 matrix, and then compute the mean of each row. This gives us a new vector 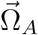 with 720 entries, which represents the average occurrence of each word in the EEGs from our control group A.

Next, we plot the vector 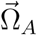 and use it as a watermark to compare against each 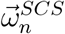 with n = 1, …, 15, which corresponds to our single case study group. For example, we plot the watermark 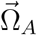 and one of the single case study vectors 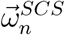 as shown in Figure 9.

**Figure 9:**
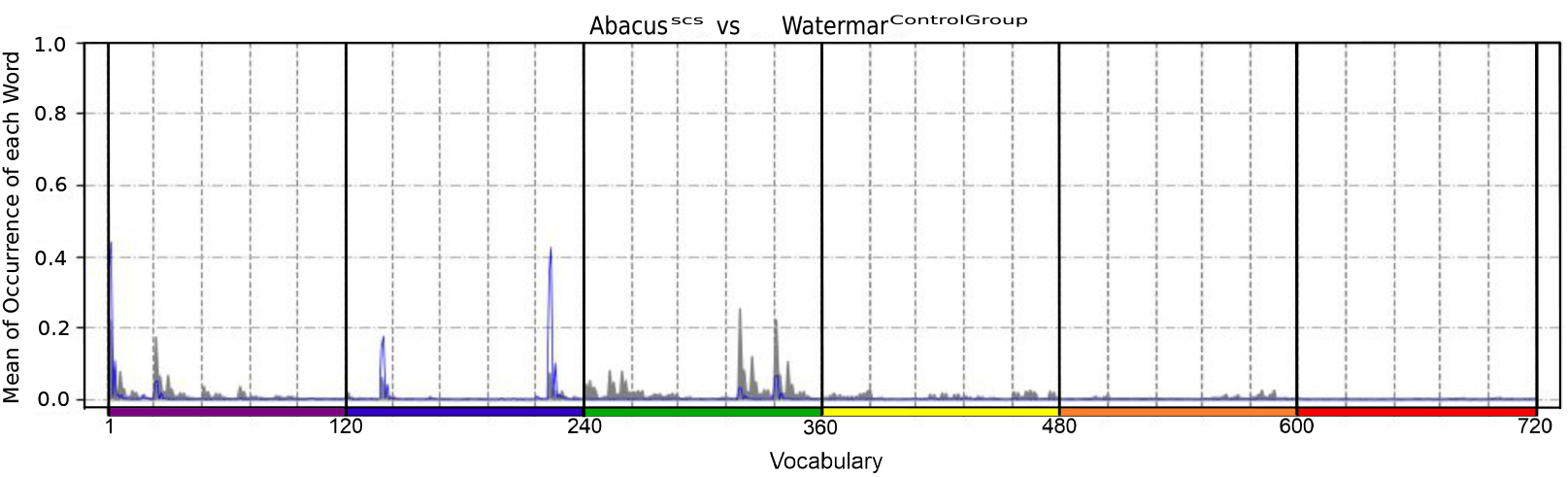
The single case study is represented in blue, while the control group watermark is displayed in gray. The colored segments below the X-axis indicate changes in the highest energy or dominant power among the ordinal ranks of the words in our vocabulary. Additionally, the vertical dashed lines represent shifts in the second-highest energy or subdominant power in the ordinal ranks of the words. This visualization mirrors the process of visual perception employed by medical experts when interpreting and reporting EEG findings.

In this way, it is possible to contrast two pieces of information and to reproduce the heuristic knowledge of descriptive statistics-epidemiological comparison done by clinical physicians supported by their accumulated expertise in reading EEGs for years. The development of this tool is left for future work.

### 2.4. Result analyses

#### 2.4.1. Machine learning algorithms

Machine learning algorithms play a crucial role in uncovering intricate patterns within data, enabling the creation of predictive models with increasing accuracy as more data is available for training. In our pursuit of developing machine learning models for disease classification, we have explored both traditional and ensemble learning algorithms following the work in [6]. In total, we have created six ML classifiers: Logistic Regression (LR), Support Vector Machine (SVM), Decision Tree (DT), Random Forest (RF), AdaBoost (AB), Gradient Boosting (GB). LR and SVM are Traditional ML models, DT and RF are Bagging models, finally AB and GB are Boosting models [38].

1. Logistic Regression (LR) LR is a statistical and machine learning technique used for binary classifications task, where the goal is to predict a binary outcome (CONTROL or SCS) based on values of input variable. Logistic Regression has the following advantages: simplicity of implementation, computational efficiency, efficiency from training perpective, ease of regularization. No scaling is required for input features.
2. Support Vector Machine (SVM) SVM can handle both classification and regression problems. In this method hyperplane needs to be defined which is the decision boundary. When there are a set of objects belonging to different classes then decision plane is needed to separate them. The objects may or may not be linearly separable in which case complex mathematical functions called kernels are needed to separate the objects which are members of different classes. SVM aims at correctly classifying the objects based on examples in the training data set. Following are the advantages of SVM : it can handle both semi structured and structured data, it can handle complex function if the appropriate kernel function can be derived. As generalization is adopted in SVM so there is less probability of over fitting. It can scale up with high dimensional data. It does not get stuck in local optima.
3. Decision Tree (DT) DT is a Supervised Machine Learning approach to solve classification and regression problems by continuously splitting data based on a certain parameter. The decisions are in the leaves and the data is split in the nodes. Decision Tree might encounter the problem of over-fitting for which Random Forest is the solution which is based on ensemble modeling approach.
4. Random Forest (RF) RF is a tree-based ensemble with each tree depending on a collection of random variables. RF can be used for both categorical response variables (Classification) and continuous response (Regression). Random forests have many advantages: they can handle Regression and multiclass classification, are faster in training and predictions, uses very few (one or two) parameters to tune, easy to implement, and can be directly used for high-dimensional problems.
5. Adaptative boosting (AB) AB, is a machine learning ensemble technique used primarily for classification problems. AB begins by fitting a classifier on the original dataset and then fits additional copies of the classifier on the same dataset but where the weights of incorrectly classified instances are adjusted such that subsequent classifiers focus more on difficult cases.
6. Gradient Boosting (GB) Gradient Boosting is another powerful ensemble machine learning technique that is used primarily for regression and classification problems. It is a generalization of AdaBoost and builds a strong predictive model by combining the predictions of multiple weaker models (typically decision trees) sequentially. Gradient Boosting, in particular, focuses on reducing the errors made by the previous models in the ensemble.

#### 2.4.2. Principal component analysis (PCA)

Principal component analysis (PCA) is a well-established method for feature extraction and dimensionality reduction. In PCA, we seek to represent the d-dimensional data in a lower-dimensional space. This will reduce the degrees of freedom; reduce the space and time complexities. The objective is to represent data in a space that best expresses the variation in a sum-squared error sense. The basic approach in principal components is theoretically rather simple. First, the d-dimensional mean vector *µ* and *d*× *d* covariance matrix *σ* are computed for the full data set. Next, the eigenvectors and eigenvalues are computed, and sorted according to decreasing eigenvalue. Call these eigenvectors e_1_ with eigenvalue λ_1_, e_2_ with eigenvalue λ_2_, and so on. Sub-sequently, the largest k such eigenvectors are chosen. In practice, this is done by looking at a spectrum of eigenvectors. It can be show that this representation minimizes a squared error criterion [42].

#### 2.4.3. Proposed methodology

We have three sets of data, the average frequencies of occurrences of each word in all channels, 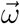 the average vocabulary used by the EEG, 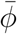, and the matrix H, which recovers the distribution of the sizes of the percentage energy per band of the corresponding three most prevalent words of the corresponding EEG channel. The statistical analysis carried out for the proposed method consists of (see Figure 10):

**Figure 10:**
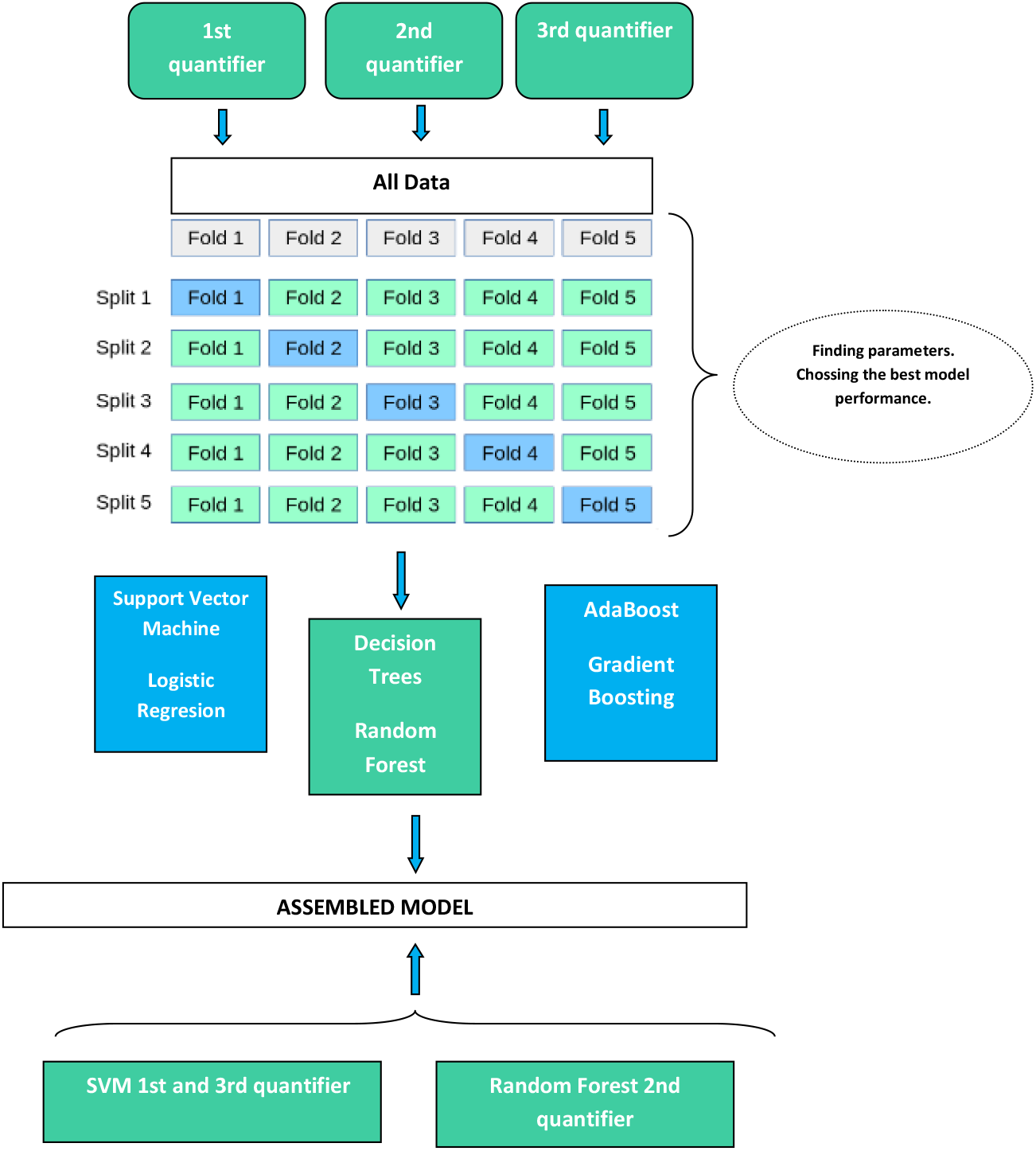
Experimental designed of the proposed model

- Application of the method to predict SCS groups from a healthy control group (CONTROL).
- A comparative study between the 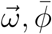 H quantifiers on six ML classifiers.
- A construction of a classifier by assembling the best-performing classifier for each quantifier through a majority vote.

#### Preprocessing data

For the matrix H we reorder the data in a new dataset so that for each individual there is a single row in the new base. The columns will be represented by the combination of the channels with each word that appears in H and will be completed with its prevalence. In addition, there will be a column for each energy band and channel whose value will be the average of the energy in the three words. more prevalent. This gives us a new base of size 45 x 921. We will refer to this new base as H as well.

We standardize the data on H with **StandarScaler()** library of python. After that we apply PCA to H and to the data frame with the observations of quantifier 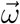 to reduce the dimensionality of the data and increase its interpretability, using **PCA()** function of python (see Figures A.11a-A.11b) **Cross-validation**.

Generalization of ML models is very important in learning-based models and to make a model that we developed to be independent of the subjects’ data we have implemented the k-fold cross-validation method by taking the k value to be 5. The 5-fold cross-validation helps to validate the ML models that we developed and helps to generalize them for the other new data as well.

#### Model building

This section describe the six machine learning models we built for the classification, with the various parameters on each model and for each of the three quantifiers 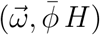. We also introduced the final model, which was constructed by assembling the best-performing classifier for each quantifier through a majority vote. These models are executed with the help of the Scikit-learn library. We have used five metrics to evaluate our developed classifier: accuracy, sensitivity, specificity, F1-Score and the “Area Under the Curve” (AUC). Each of them is discussed below.

True Positive (TP): it refers to a situation where a model correctly identifies or classifies a positive condition when the condition is actually present. True Negative (TN): it refers to a situation where a model correctly identifies or classifies a negative condition when the condition is actually absent. False Positive (FP): It refers to a situation where a test or model incorrectly identifies or classifies a negative condition as positive, when in reality, the condition is not present

False Negative (FN): It refers to a situation where a test or model incorrectly identifies or classifies a positive condition as negative, when in reality, the condition is present.

Accuracy: The overall accuracy of the model is given by a formula

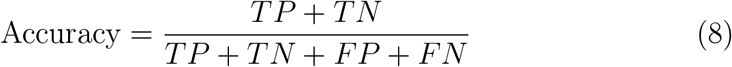

Sensitivity (true positive rate): It measures the ability of the model or test to correctly identify positive instances or cases when they are truly positive.

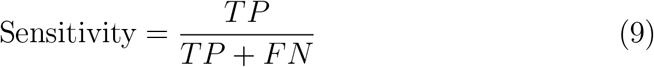

Specificity: It measures the ability of the model or test to correctly identify negative instances or cases when they are truly negative.

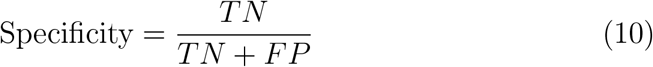

F1-Score: is a single metric that combines both Precision and Recall (Sensitivity) into a single value. It is calculated as the harmonic mean of Precision 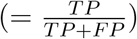 and Recall.

The formula for the F1-score is:

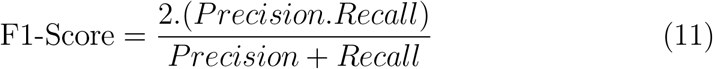

The “Area Under the Curve” (AUC) quantifies the performance of a model in distinguishing between positive and negative classes across different decision thresholds. In practical terms, a higher AUC indicates a better-performing model, particularly in terms of its ability to discriminate between positive and negative cases The details of the six classifiers, for which we conducted parameter tuning within a specified range, are provided in tables 1-3, showcasing the highest recall, accuracy, and AUC achieved.

**Table 1:** 1st Quatifier: 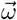.

| Classifier | Parameters |
| --- | --- |
| Support Vector Machine | Kernel='linear'; gamma=0.1; C=100; random-state=0 |
| Logistic Regression | solver='lbfgs'; C=100 |
| Decision Tree | max-depth=5 |
| Random Forest | n-estimators=100; max-depth=20; random-state=0 |
| AdaBoost | n-estimators=20; learning-rate=0.1 |
| Gradient Boost | n-estimators=20; max-dept=9 |

**Table 2:** 2nd Quatifier: 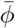.

| Classifier | Parameters |
| --- | --- |
| Support Vector Machine | Kernel='sigmoid'; gamma=0.1; C=0.1; random-state=0 |
| Logistic Regression | solver='liblinear'; C=10 |
| Decision Tree | max-depth=3 |
| Random Forest | n-estimators=50; criterion="entropy"; max leaf nodes=2 ; random-state=0 |
| AdaBoost | n-estimators=20; learning-rate=0.01 |
| Gradient Boost | n-estimators=20; max-dept=3 |

**Table 3:** 3rd Quatifier: *H*.

| Classifier | Parameters |
| --- | --- |
| Support Vector Machine | Kernel='sigmoid'; gamma=0.1; C=1; random-state=0 |
| Logistic Regression | solver='sag'; C=0.1 |
| Decision Tree | max-depth=5 |
| Random Forest | n-estimators=200; max-depth=30; random-state=0 |
| AdaBoost | n-estimators=20; learning-rate=0.01 |
| Gradient Boost | n-estimators=20; max-dept=5 |

## 3. Results

We studied the quantifiers 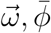 *H* on six AI models. We have recorded various performance metrics like accuracy, sensitivity, specificity, F1-Score and AUC, which are presented in table 4. The performance parameters of six models are illustrated in the figures A.12-A.16. We refers as 1st quantifier, 2nd quantifier and 3rd quantifier on the graphics to the quantifiers 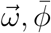 and *H* respectively. Based on the results obtained from the various metrics, we selected SVM models for the first and third quantifiers, while we opted for a Random Forest model for the second quantifier. Details of the parameters used are detailed in the tables 1-3. We combined the three models through a majority voting process. The metrics resulting from this assembly model are presented in the table 5. We have calculated the confusion matrix for the SVM algorithm apply to the the first and third quantifiers, the Random Forest algorithm for the second quantifier, and the assembly model.

**Table 4:** Results of the metrics obtained on the six Machine learning algoritms.

|  | Accuracy (%) |  |  | Sensitivity (%) |  |  | Specificity (%) |  |  | F1-Score |  |  | AUC |  |  |
| --- | --- | --- | --- | --- | --- | --- | --- | --- | --- | --- | --- | --- | --- | --- | --- |
| | $\bar{\omega}$ | $\bar{\phi}$ | $H$ | $\bar{\omega}$ | $\bar{\phi}$ | $H$ | $\bar{\omega}$ | $\bar{\phi}$ | $H$ | $\bar{\omega}$ | $\bar{\phi}$ | $H$ | $\bar{\omega}$ | $\bar{\phi}$ | $H$ |
| SVM | 93.33 | 86.67 | 88.89 | 93.33 | 86.67 | 86.67 | 93.33 | 86.67 | 90 | 90.29 | 80.76 | 84.14 | 93.33 | 86.67 | 88.33 |
| LR | 88.89 | 88.89 | 88.89 | 86.67 | 86.67 | 80 | 90 | 90 | 93.33 | 84.33 | 83.62 | 80.29 | 88.33 | 88.33 | 86.67 |
| GB | 80 | 86.67 | 75.56 | 66.67 | 73.33 | 60 | 86.67 | 93.33 | 83.33 | 67.9 | 76.48 | 61 | 76.67 | 83.33 | 71.67 |
| RF | 86.67 | 93.33 | 88.89 | 60 | 86.67 | 73.33 | 100 | 96.67 | 96.67 | 72 | 89.33 | 79.14 | 80 | 91.67 | 85 |
| AB | 91.11 | 93.33 | 75.56 | 80 | 86.67 | 60 | 96.67 | 96.67 | 83.33 | 85.33 | 89.33 | 61 | 88.33 | 91.67 | 71.67 |
| DT | 80 | 86.67 | 68.89 | 66.67 | 73.33 | 60 | 86.67 | 93.33 | 73.33 | 67.67 | 76.48 | 57 | 76.67 | 83.33 | 66.67 |

**Table 5:** Results of the metrics obtained on the Assembly Model and the best models for each quantifier.

|  | Accuracy (%) | Sensitivity (%) | Specificity (%) | F1-Score (%) | AUC (%) |
| --- | --- | --- | --- | --- | --- |
| Assembly Model | 95.55 | 93.33 | 96.67 | 93.14 | 95.00 |
| 1st Quantifier $\bar{\omega}$ | 93.33 | 93.33 | 93.33 | 90.29 | 93.33 |
| 2nd Quantifier $\bar{\phi}$ | 93.33 | 86.67 | 96.67 | 89.33 | 91.67 |
| 3rd Quantifier $H$ | 88.89 | 86.67 | 90.00 | 84.14 | 88.33 |

## 4. Discussion

Generally, signal processing methods separate the information by bands, [4]. We suggests an alternative method for analyzing EEG signals by examining the behavior of the accumulated power of all bands simultaneously, which could be more efficient than separating the information by bands.

Artifacts in EEG signals during prolonged studies produce different types of noise, including brief, intermittent, and single-channel artifacts, among others. These artifacts pose a challenge for neurophysiological diagnosis. In [45], a method was developed to detect discharges even in high noise environments. Our method describes the behavior of recording energy by using averaged values in the calculation of various metrics. This averaging process helps to smooth out some of the artifacts that interfere with the descriptive neurophysiological diagnosis of cumulative power.

On the other hand, much of the older equipment used in medical practice, such as in our case, has a low sampling rate. This poses a problem for the detection of spikes in intercritical recordings for focal epilepsies [5]. In [14], colleagues present a method for automatically detecting epileptic patterns in EEG using deep learning. The multi-head self-attention mechanism effectively identifies long-term dependencies as well as dynamic temporal correlations between short-term temporal pattern features and sequential relationships. Contextual representations are introduced into a bidirectional short-term memory network (bilstm), achieving a sensitivity of 96.5 %, specificity of 97.04%, F1-score of 96.6%, and precision of 96.2%.

While these results surpass ours, our method focuses on the study of basal activity (wakefulness with closed eyes) in interictal recordings, providing insights for both diagnosis and monitoring the progression of patients already diagnosed with epilepsy and/or other clinical conditions through quantifiable metrics. The method we propose demonstrates how the behavior of energy ratios in the background activity can be quantified and used to supplement evolutionary neurophysiological diagnosis in epilepsies in general and in other clinical situations.

Overall, the proposed method could be a valuable addition to the existing tools and techniques for analyzing EEG signals, and it could potentially lead to more accurate and efficient diagnosis and treatment of neurological and neuropsychiatric conditions [22].

## 5. Conclusions

The functional neurophysiological diagnosis of an EEG is a multidimensional process that involves evaluating various characteristics to determine whether the recording is normal [58, 20]. One critical aspect is the energy distribution, which we quantify using specific metrics to provide a structured, objective analysis.

Our method successfully classifies 96.6% of control group patients as normal and detects abnormalities even for individuals without specialized training in clinical neurophysiology. A key strength of this approach is its reliance on a rigorously defined control group, ensuring diagnostic consistency even when the underlying clinical pathology is unknown. The proposed metrics demonstrate stable performance, enabling different laboratories to establish their own control groups and seamlessly integrate the method into daily clinical practice.

The methodology developed in this study is robust in both information encoding and systematic EEG interpretation. By standardizing the assessment of energy distribution, these metrics improve the objectivity of EEG evaluation across different experimental designs and hardware configurations. Given the substantial variability in medical EEG reporting, as highlighted by a recent study analyzing seizure onset patterns in 1799 EEG recordings, our method addresses the need for quantitative, standardized diagnostic descriptors.

Beyond classification accuracy, the proposed quantification approach enhances inter-rater reliability and lays the foundation for automated diagnostic tools. Importantly, it distinguishes neurophysiological descriptive diagnosis from psychiatric or neurological diagnoses, ensuring that EEG interpretation remains within its intended clinical scope as a complementary diagnostic tool. The method’s versatility allows EEG analysis to be independent of hardware constraints, supporting various channel configurations. Additionally, its computational simplicity makes it suitable for real-world clinical applications, though further refinements may facilitate broader adoption.

In conclusion, this FFT-based quantification framework represents a significant step toward standardizing EEG diagnostics. By improving objectivity and reproducibility while maintaining clinical relevance, it contributes to a more accessible and structured approach to neurophysiological assessment.

## Author Contributions

Conceptualization, Juan Manuel Diaz López and Carina Boyallian; methodology, Juan Manuel Díaz López; software, Juan Manuel Díaz López and José Curetti; validation, Juan Manuel Díaz López and Carina Boyallian and Vanesa Meinardi.; formal analysis, Juan Manuel Díaz López and Carina Boyallian; investigation, Juan Manuel Díaz López and Carina Boyallian; resources, Juan Manuel Díaz López; EEG control group data collection, Juan Manuel Díaz López; data curation, Juan Manuel Díaz López and Vanesa Meinardi.; writing—original draft preparation, Juan Manuel Díaz López, Carina Boyallian and Vanesa Meinardi; writing—review and editing, Juan Manuel Díaz López and Carina Boyallian; visualization, Juan Manuel Díaz López and Vanesa Meinardi.; supervision, Carina Boyallian and Hugo Díaz Farjreldines. All authors have read and agreed to the published version of the manuscript.

## Funding

This research received no external funding.

### Institutional Review Board Statement

The study was conducted in accordance with the Declaration of Helsinki, and approved by the Ethics Committee of Hospital Córdoba: Comite Institucional de Etica de la investigación en salud del adulto. Ministerio de Salud de la Provincia de Córdoba. (protocol code CO000152 and 20-03-2017).

### Informed Consent Statement

Informed consent was obtained from all subjects involved in the study.

## Data Availability Statement

The data presented in this study are available on request from the corresponding author.

## Acknowledgments

The first author thanks Dr. Graciela Lucatelli and Dr. Atilio Bollo, for their valuable support. The last author thanks Dr. Lamberti, Dr. Buonanote and Dr. Rosso Osvaldo A. for useful discussions.

## Conflicts of Interest

The authors declare no conflicts of interest.

## Appendix A

**Figure A.11:**
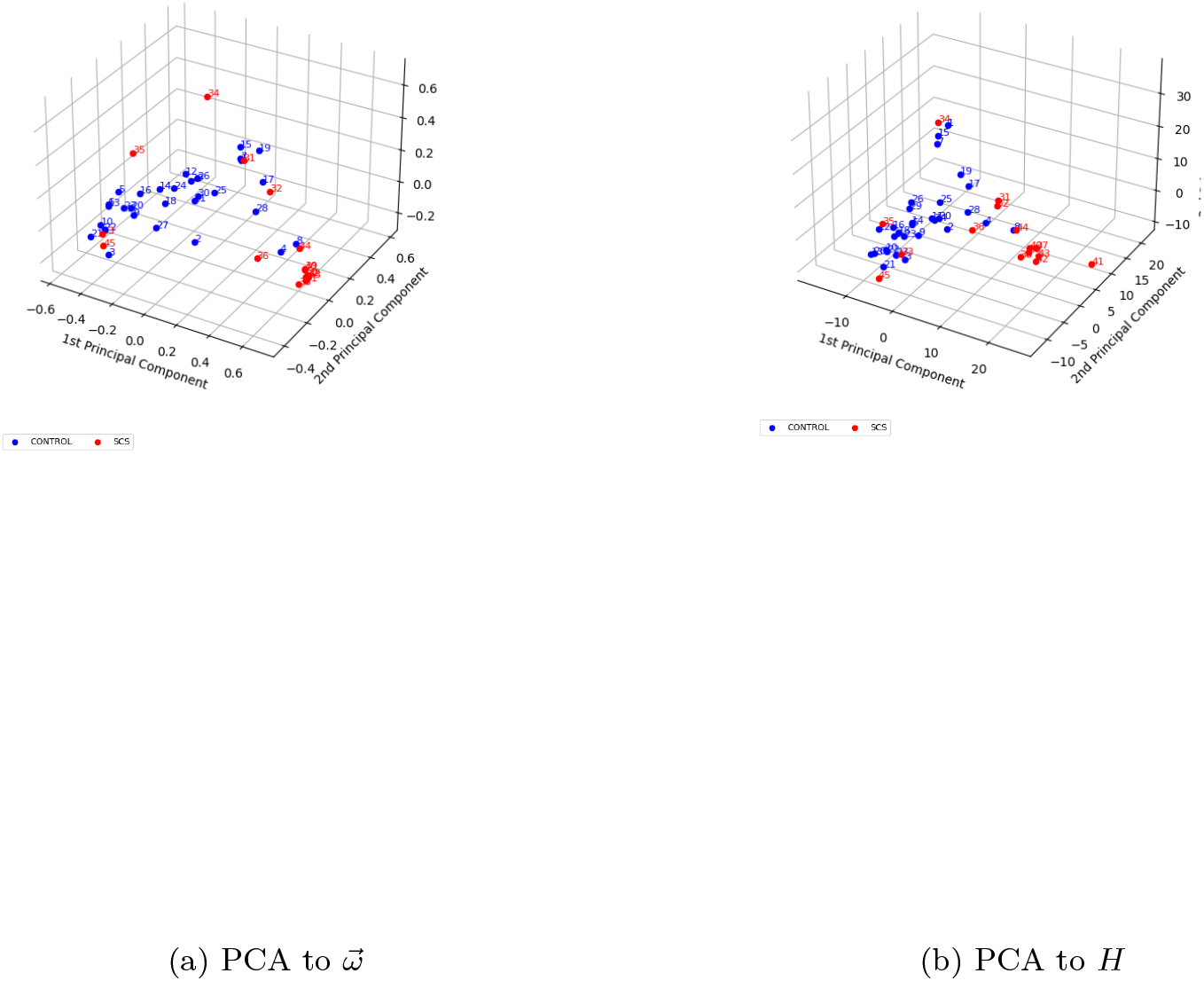
Principal Component Analysis

**Figure A.12:**
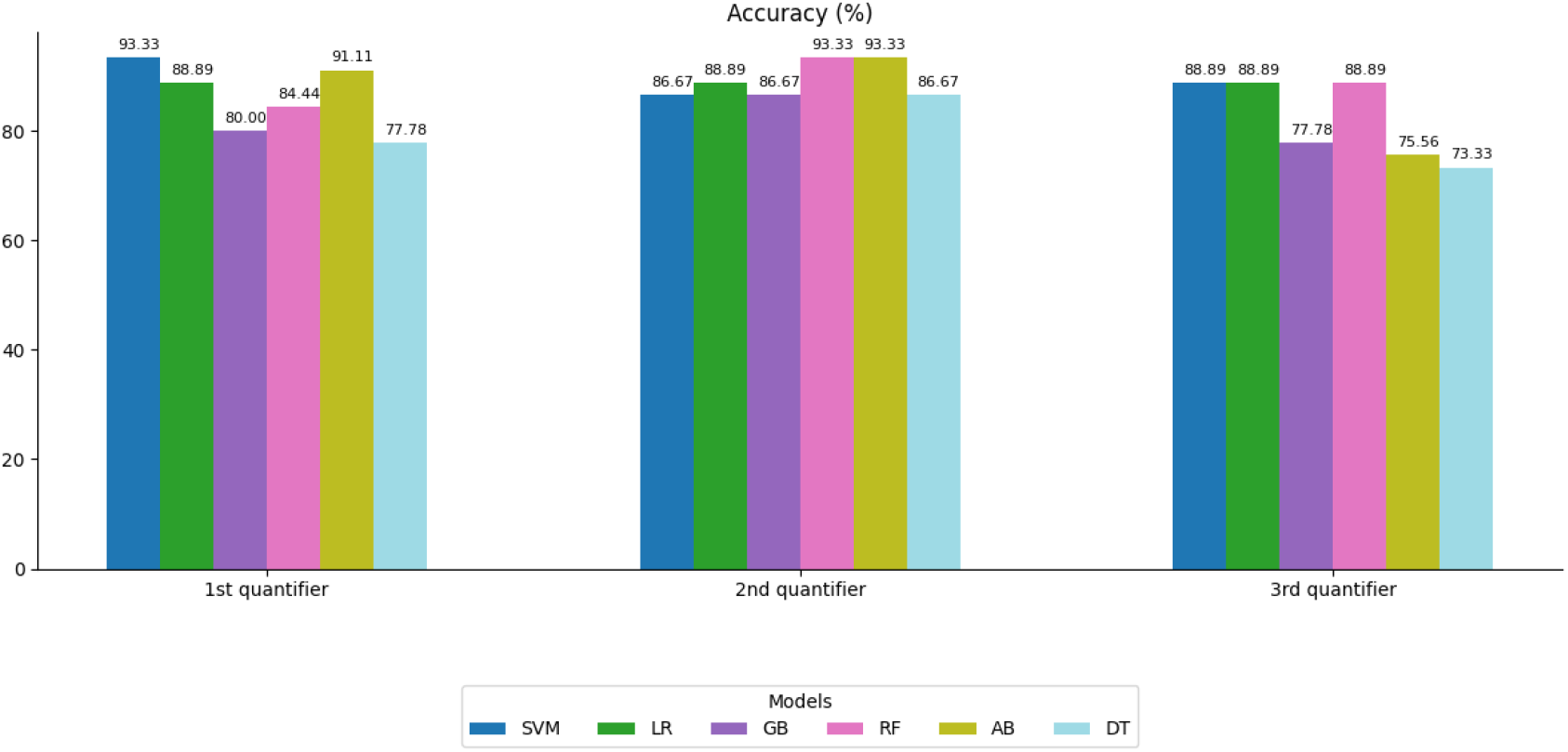
Accuracy (%) of six classifiers under three quantifiers

**Figure A.13:**
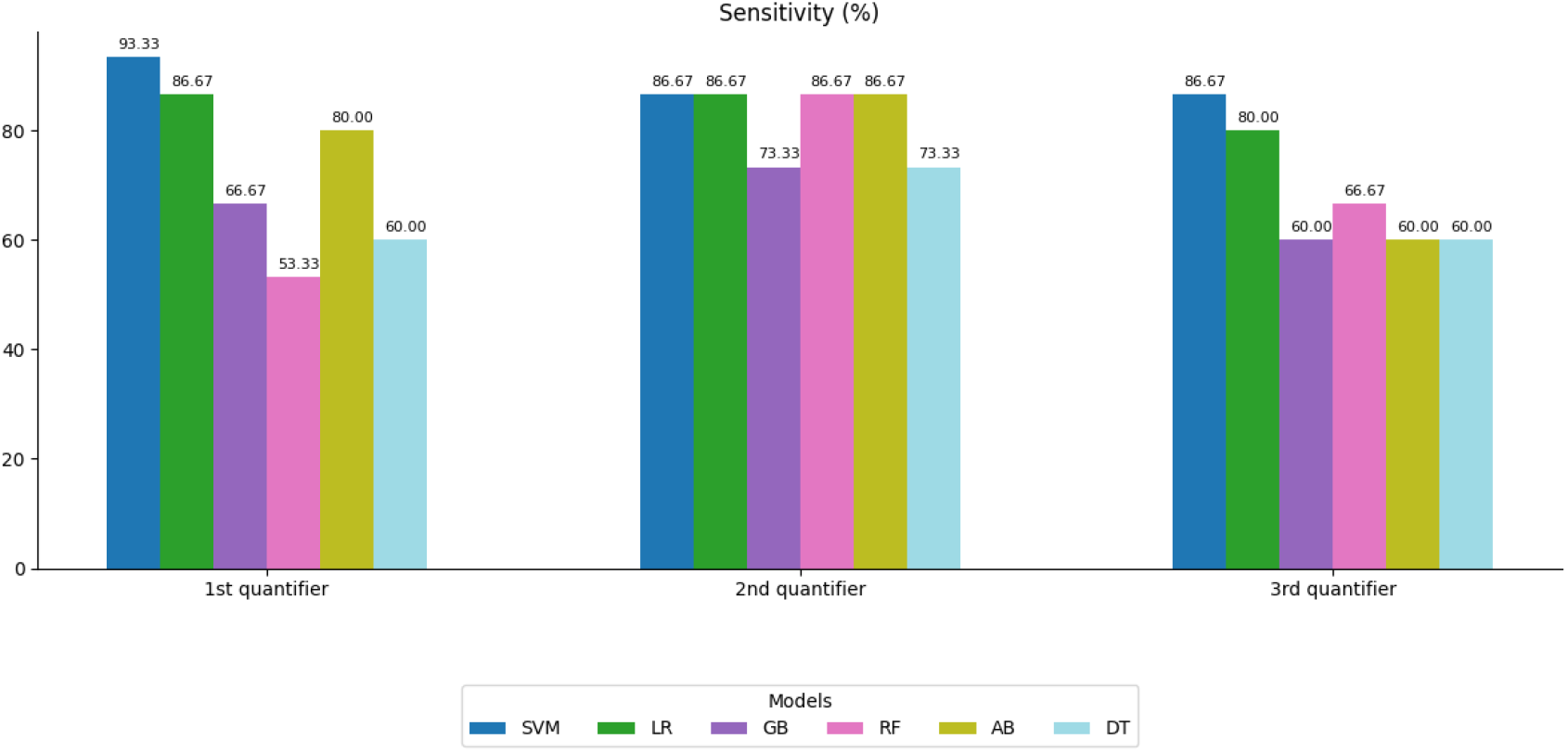
Sensitivity (%) of six classifiers under three quantifiers

**Figure A.14:**
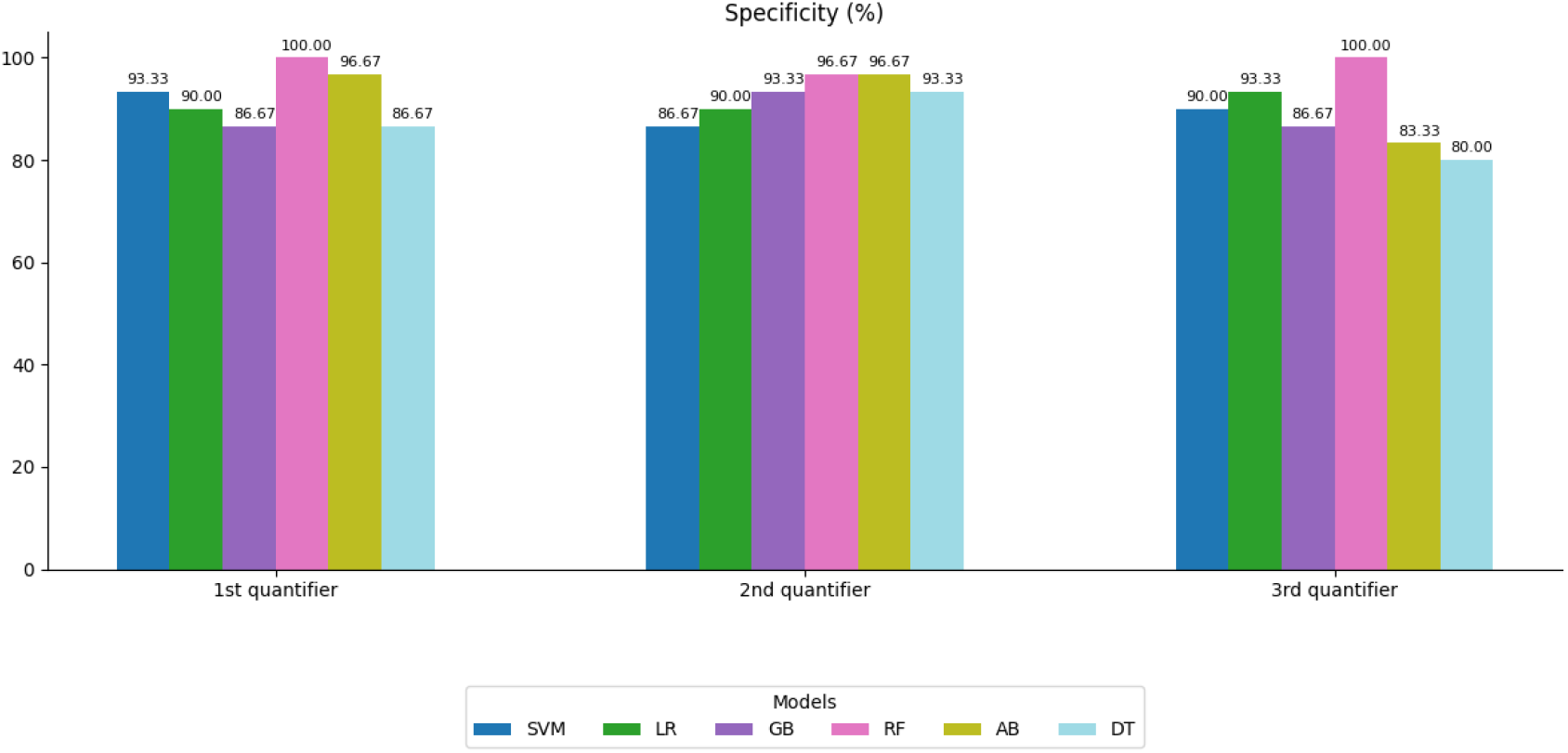
Specificity (%) of six classifiers under three quantifiers

**Figure A.15:**
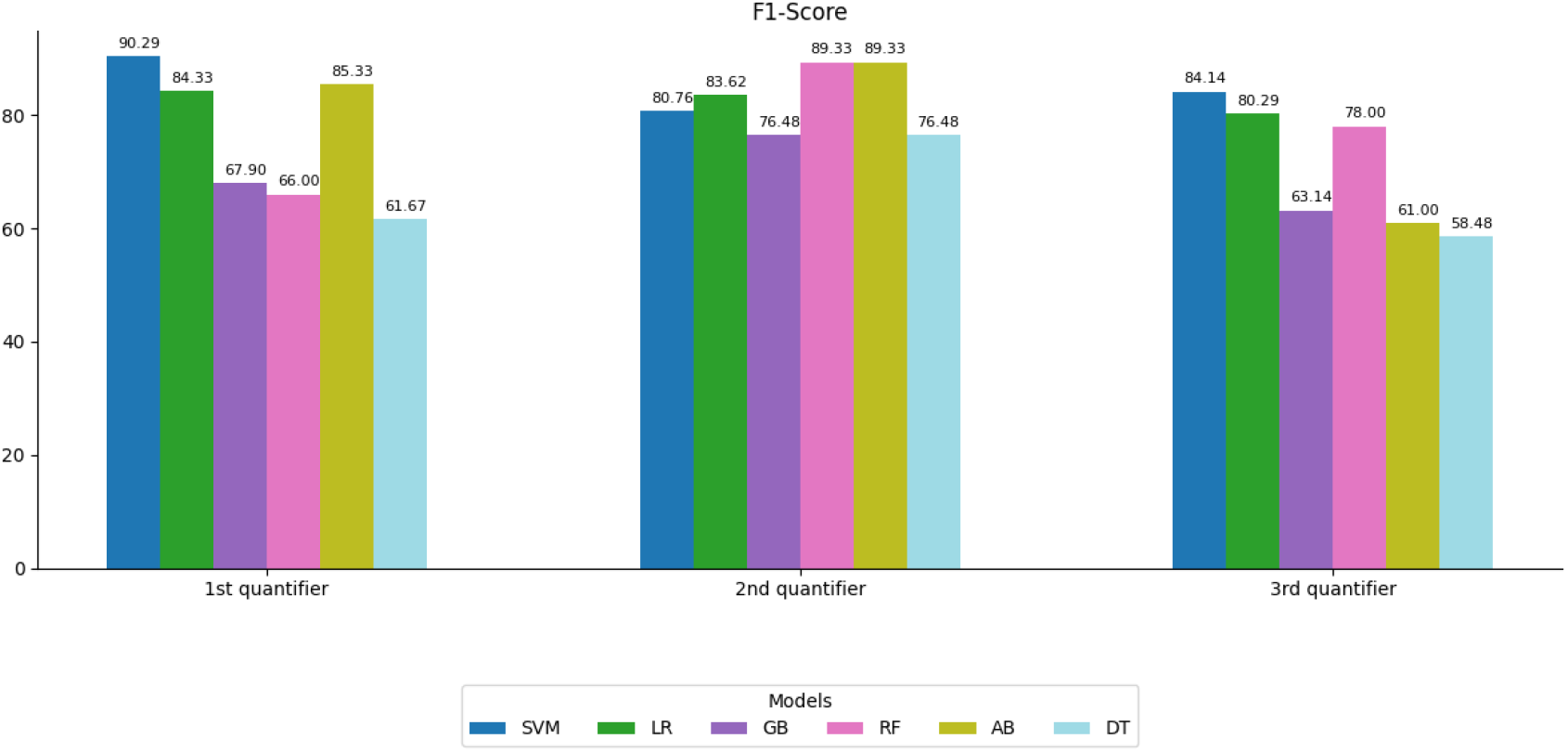
F1-Score (%) of six classifiers under three quantifiers

**Figure A.16:**
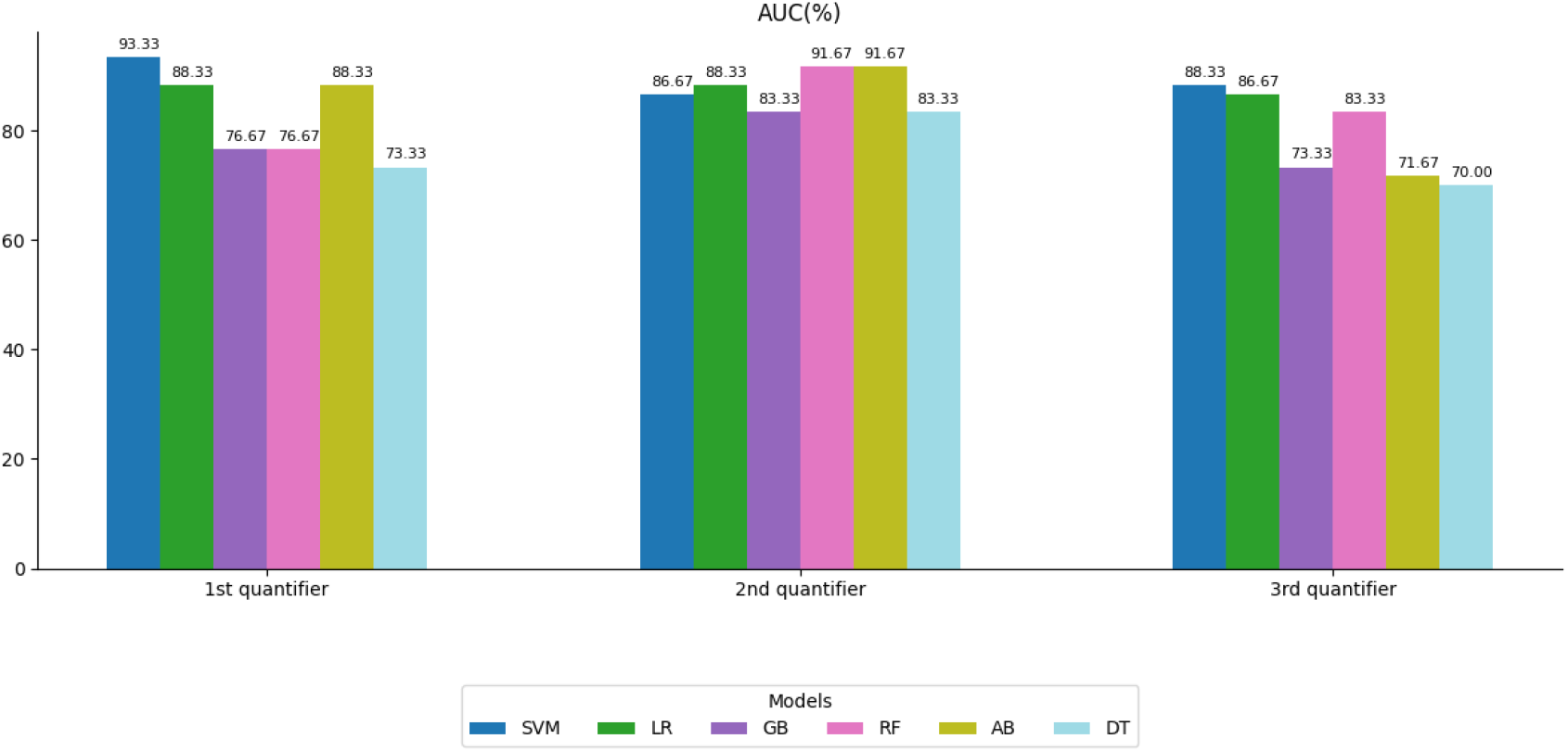
AUC (%) of six classifiers under three quantifiers

**Table A.6:**
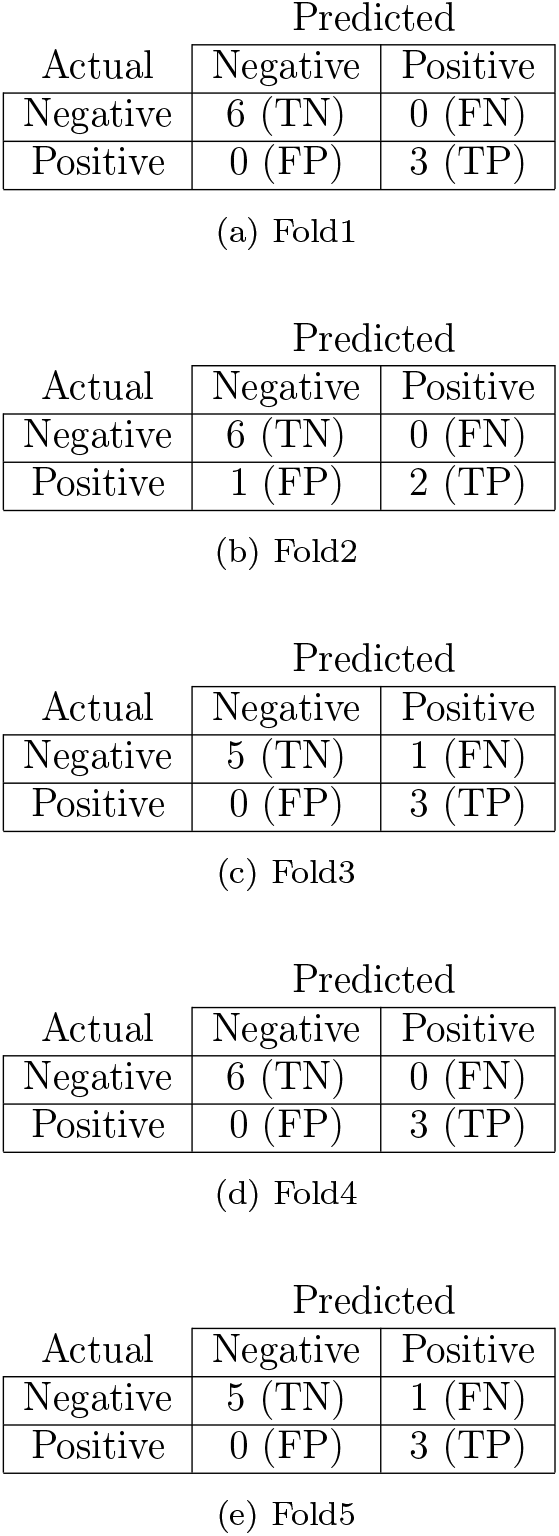
Confusion Matrices for the SVM model applied to the 1st quantifier.

**Table A.7:** Confusion Matrices for the Random Forest model applied to the 2nd quantifier.

(a) Fold1
| Actual | Predicted |  |
| --- | --- | --- |
|  | Negative | Positive |
| Negative | 6 (TN) | 0 (FN) |
| Positive | 2 (FP) | 1 (TP) |

(c) Fold3
| Actual | Predicted |  |
| --- | --- | --- |
|  | Negative | Positive |
| Negative | 5 (TN) | 1 (FN) |
| Positive | 1 (FP) | 2 (TP) |

(d) Fold4

**Table A.8:** Confusion Matrices for the SVM model on the 3rd quantifier.

(b) Fold2
| Actual | Predicted |  |
| --- | --- | --- |
|  | Negative | Positive |
| Negative | 4 (TN) | 2 (FN) |
| Positive | 0 (FP) | 3 (TP) |

(c) Fold3

**Table A.9:**
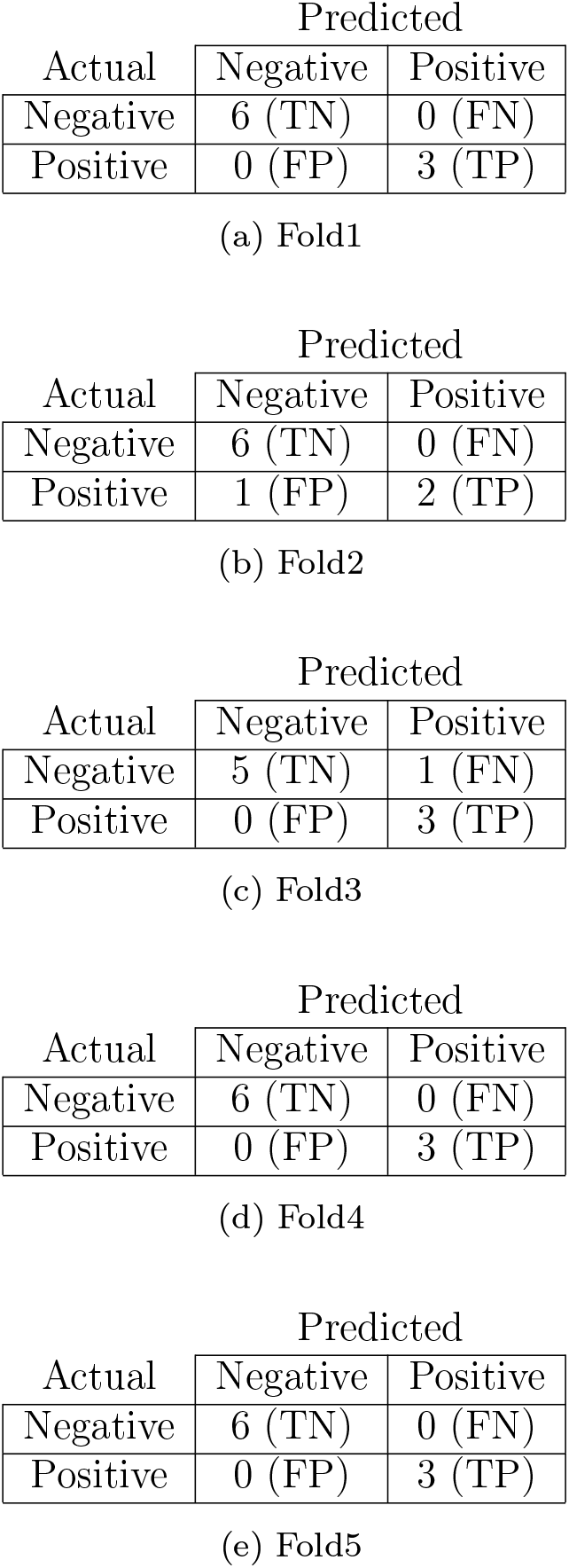
Confusion Matrices for the assembly of SVM and Random Forest models.

## Notes

### Competing Interest Statement

The authors have declared no competing interest.

### Author Declarations

The research protocol was approved by the ethics committee of Hospital Cordoba, Cordoba, Argentina and registered in the Ministry of Health of the Cordoba Province on 20-03-2017 under number 3170. (protocol code CO000152 and 20-03-2017).

